# Toward Understanding COVID-19 Pneumonia: A Deep-learning-based Approach for Severity Analysis and Monitoring the Disease

**DOI:** 10.1101/2020.11.24.20235887

**Authors:** Mohammadreza Zandehshahvar, Marly van Assen, Hossein Maleki, Yashar Kiarashi, Carlo N. De Cecco, Ali Adibi

**Affiliations:** School of Electrical and Computer Engineering, Georgia Institute of Technology, Atlanta, GA, USA; Department of Radiology and Imaging Sciences, Emory University School of Medicine, Atlanta, GA, USA

**Keywords:** deep learning, interpretable models, dimensionality reduction, X-ray, COVID-19, severity analysis, monitoring progression

## Abstract

We report a new approach using artificial intelligence to study and classify the severity of COVID-19 using 1208 chest X-rays (CXRs) of 396 COVID-19 patients obtained through the course of disease at Emory Healthcare affiliated hospitals (Atlanta, GA, USA). Using a two-stage transfer learning technique to train a convolutional neural network (CNN), we show that the algorithm is able to classify four classes of disease severity (normal, mild, moderate, and severe) with average area under curve (AUC) of 0.93. In addition, we show that the outputs of different layers of the CNN under dominant filters provide valuable insight about the subtle patterns in the CXRs, which can improve the accuracy in the reading of CXRs by a radiologist. Finally, we show that our approach can be used for studying the disease progression in single patients and its influencing factors. The results suggest that our technique can form the foundation of a more concrete clinical model to predict the evolution of COVID-19 severity and the efficacy of different treatments for each patient through using CXRs and clinical data in early stages. This will be essential in dealing with the upcoming waves of COVID-19 and optimizing resource allocation and treatment.

## 1 Introduction

COVID-19 was declared a global health emergency by the World Health Organization in January of 2020, and governments have put unprecedented measures to halt the transmission of the virus [1]. However, the healthcare systems are still struggling with the massive influx of patients. The virus mostly emerges with mild or no symptoms in the early stages. However, it can rapidly cause pneumonia and lung opacity in patients resulting in a fatality or long-term damages to the lung [2–4]. This emphasizes the importance of timely diagnosis and evaluation of the severity degree and other features of the disease to optimize resource allocation for extensive treatments such as intensive care unit (ICU).

From the beginning of the pandemic, researchers started to develop different platforms and test kits for diagnosis of COVID-19. Prior to the availability of reverse-transcription polymerase-chain-reaction (RT-PCR) tests, chest X-rays (CXRs) and computed tomography (CT) scans were used to diagnose COVID-19 [5, 6]. Chest imaging, CT in particular, shows a characteristic manifestation of COVID-19, even in patients with limited symptoms. However, this use of CT places a considerable burden on radiology departments and poses an immense challenge for infection control. There have also been many cases with positive COVID-19 without any pulmonary manifestations [7]. The significant efforts and advances to foster the development of cheaper, more accurate, faster, and easier-to-use test kits and the aforementioned reasons raise questions about using medical imaging as a tool for detecting the disease. Instead, these modalities can play a crucial role in determining the severity degree of the disease and understanding the dynamics of its development from mild to severe in different patients as well as predicting its evolution and assessing the efficacy of different treatments[2, 3, 8–14], which are not feasible in RT-PCR and other laboratory test kits.

Predicting the severity degree of COVID-19 and its impacts on the lung are of great importance, enabling monitoring the progress of the disease over time and helping resource allocation in hospitals [15–17]. Several studies have shown that the severity classification and severity progression of COVID-19 are highly related to ICU admission, length of hospital stay, and optimal planning of follow-ups [6, 8, 10, 12, 14]. Despite being less sensitive than chest CT for diagnosis of COVID-19-related pneumonia, especially in the early phases of the disease, CXRs are often being utilized in the first-line assessment of patients due to their affordability and accessibility. In its recent guideline, the American College of Radiography (ACR) recommends portable CXR in ambulatory care facilities over CTs [18]. The British Medical Journal (BMJ) Best Practice recommendations for COVID-19 management also endorses the use of CXR in all patients with suspected pneumonia [19]. A retrospective study by Wong H.Y.F. et al. [4] on 64 patients with an RT-PCR-confirmed COVID-19 diagnosis found that the most common COVID-19-related signs on CXRs were consolidation (47%) and ground-glass opacities (33%). These alterations mostly had a bilateral, peripheral, lower zone distribution and were rarely associated with pleural effusion. In their study, 69% of patients had CXR abnormalities on the baseline X-ray, and another 11% developed CXR alterations later on in the study. As a result, CXR can be a very effective modality for monitoring the progression of the disease and its effect on the pulmonary system. However, the increasing number of patients and the large number of CXRs burden an unprecedented workload on radiologists and calls for automatic severity prediction and monitoring systems more than any time.

Artificial intelligence (AI) may be a viable solution for the automatic diagnosis and prognosis of COVID-19 and unburdening physicians and radiologists of the high workload. Recent studies in using AI for diagnosis of the disease from CXRs and CT scans shows the capabilities of these methods in providing automatic and rapid diagnosis [20–26]. However, despite the unique opportunities enabled by AI, most existing works focus on classification and detection of COVID-19 [27–32], rather than classifying the severity degree of the disease [33–37] and providing intuition about its potential evolution to enable a decision-making process, which can also facilitate unprecedented knowledge discovery in COVID-19. In this paper, we demonstrate an effective AI approach for this important purpose.

While most of the reported works on CXR for diagnosis of COVID-19 use online datasets that might combine CXRs from different sources and different settings, in this study, we use an authentic dataset from Emory Healthcare affiliated hospitals (Atlanta, GA, USA), which is labeled by an expert radiologist (CNDC) with 15+ years of experience. Here, we report an AI model based on training a deep convolutional neural network (CNN) for predicting the severity of COVID-19 using CXR images. We will also use interpretable models through dimensionality reduction and visualization of the outputs of different layers of the trained CNN to obtain valuable insight about the evolution of the disease as well as the decision-making process [38–43]. We show that our model can predict the severity degree of pneumonia caused by COVID-19 from CXR images with the area under the curve (AUC) of 0.97, 0.90, 0.90, and 0.95 for normal, mild, moderate, and severe COVID-19 classes over an unseen test set. We will also visualize the most important and dominant features that the CNN extracts from CXR images at each layer of the CNN by applying a pruning algorithm based on the average percentage of zeros (APoZ) [44]. Finally, we leverage the uniform manifold approximation and projection (UMAP) method [45] to form the low-dimensional manifold of the input-output relation (i.e., X-ray images to COVID-19 severity classes) to clearly monitor the disease progression.

## 2 Results

### 2.1 Emory hospital COVID-19 X-ray dataset for training and evaluation

In total, 1208 CXRs from 396 patients are used in this study. CXRs are consecutive samples from patients who had a clinically-performed positive COVID-19 RT-PCR test during the same admission as their CXR imaging at Emory-Healthcare-affiliated hospitals (Atlanta, GA, USA). An expert cardiothoracic radiologist with 15+ years of experience in reading CXRs has labeled all CXRs. These CXRs are blinded and randomized for the classification of the disease severity and the reader has had no insight into clinical data and/or outcomes. The CXRs are classified into normal, mild, moderate, and severe classes depending on the consolidation and opacity degrees (see section 4 for more details about the labeling process). The number of images are 178, 506, 384, and 140 for the normal, mild, moderate, and severe classes, respectively (see Table 1). The dataset includes 196 males and 200 females with an average age of 63.1. We use 966 CXRs for training and keep the rest for evaluating the performance of the model.

**Table 1.** Patients cohort utilized in the study. All the patients in this study are diagnosed with COVID-19 using RT-PCR test. CXRs for training and testing the model are chosen randomly from 1208 CXRs.

|  |  |  |
| --- | --- | --- |
| Total Number of Patients | 396 |  |
| Total Number of CXRs | 1208 |  |
| CXRs Class Distribution | Normal | 178 (14.7%) |
|  | Mild | 506 (41.9%) |
|  | Moderate | 384 (31.8%) |
|  | Severe | 140 (11.6%) |
| Age Statistics | Min | 18 |
|  | Mean | 63.10 |
|  | Max | >90 |
|  | Standard Deviation | 16.19 |
| Gender Distribution in CXRs | Male | 640 (53%) |
|  | Female | 568 (47%) |
| Gender Distribution in Patients | Male | 196 (49.5%) |
|  | Female | 200 (50.5%) |

### 2.2 Deep-learning (DL)-based model for classifying the severity of COVID-19

The CNN (shown in Fig. 1) is trained using 80% of the CXRs through a two-stage transfer-learning algorithm to assess the severity of COVID-19 for an input CXR (see section 4 for details)[46, 47]. The receiver operating characteristic curve (ROC) for the average performance of the DL-based model over 10 unseen CXR test sets (sampled from the dataset using the bootstrap method [48, 49]) is shown in Fig. 2a. The average AUC over 10 test sets for normal, mild, moderate, and severe classes are 0.97, 0.90, 0.90, and 0.95, respectively. The lower AUC for mild and moderate classes in comparison with normal and severe classes is in line with their larger overlap with other classes and the fairly subtle differences between these categories (i.e., mild and moderate) and surrounding categories (i.e., normal and severe). Based on the confusion matrix of the model (Fig. 2b), the average recall (i.e., sensitivity) for normal, mild, moderate, and severe classes is 0.83, 0.80, 0.68, and 0.65, respectively. According to Fig. 2b, the misclassification rate for non-adjacent classes (e.g., normal vs. moderate, severe vs. normal, etc.) is zero with the exception of 0.01 for the severe vs. mild case, which is negligible. The model specificity is 0.93, 0.81, 0.87, and 0.95 for the normal, mild, moderate, and severe classes, respectively. Similar to the AUC, the specificity of the model is lower for intermediate classes (i.e., mild and moderate) due to the reasons explained earlier.

**Figure 1.**
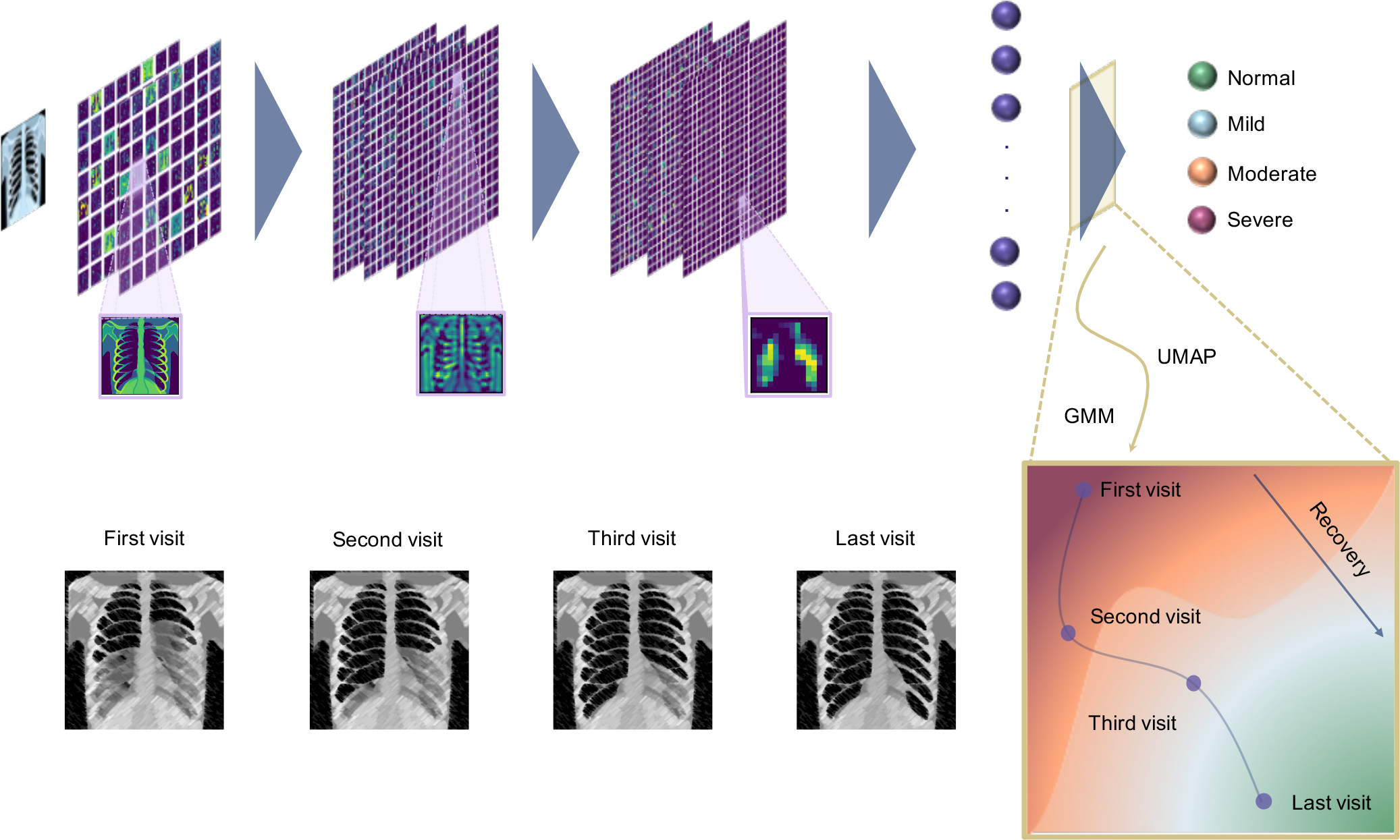
Workflow of COVID-19 severity assessment and prognosis model. The input CXR is fed into the CNN and will be processed by the network to extract the features and classify the severity degree of the disease. At each layer, the outputs of the most dominant CNN layers will be extracted and visualized using a pruning approach to understand the decision-making process. To monitor the progress of the disease, we reduce the dimensionality of the last fully connected layer using UMAP and use the Gaussian Mixture Model (GMM) to cluster the space into different severity regions. For series of input CXRs of a patient, we can visualize and monitor the progress of the disease over time.

**Figure 2.**
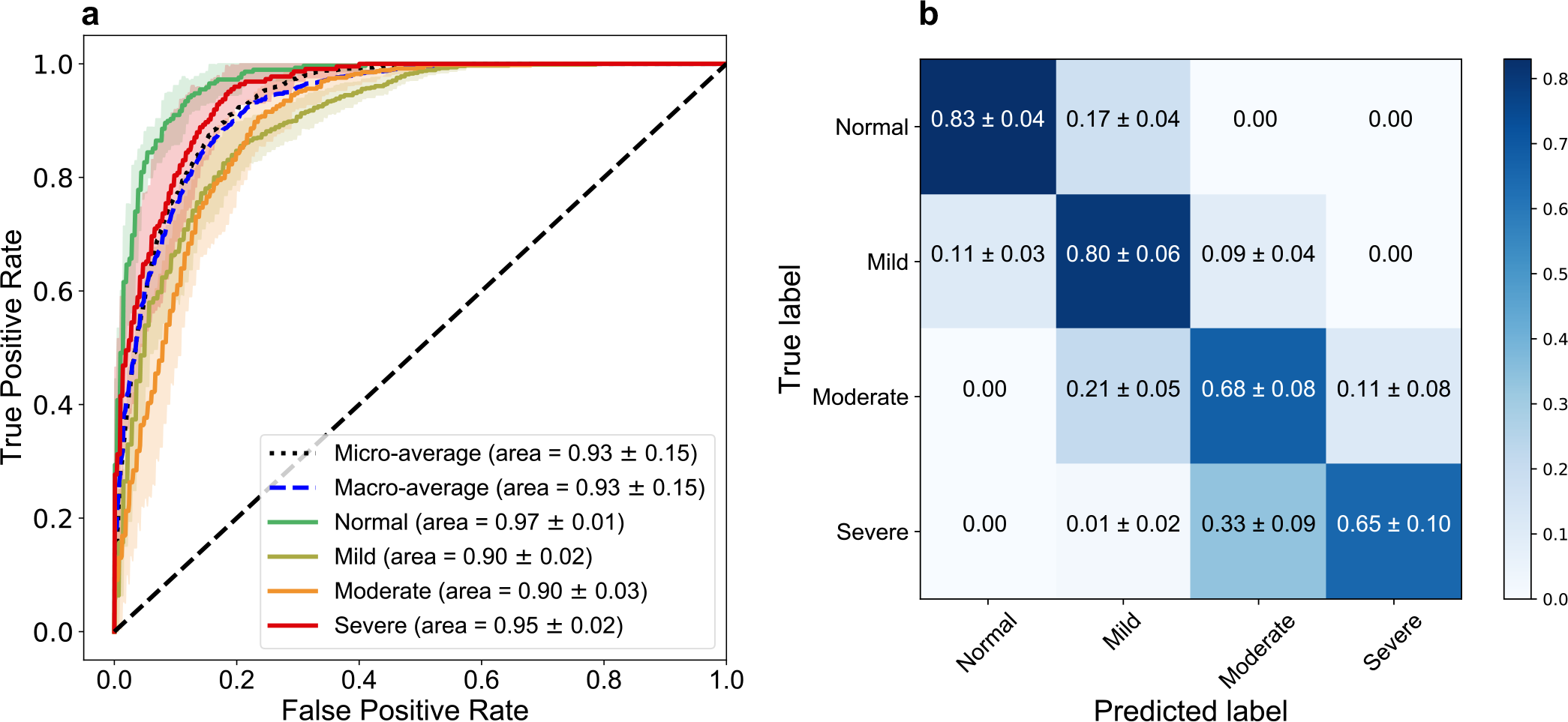
Model Performance. **a** The receiver operating characteristic (ROC) curves for normal, mild, moderate, and severe classes and the micro- and macro-average ROCs. The solid lines show the means of ROCs over 10 independent test sets, sampled using the bootstrap method. The standard deviations for the ROCs are represented as shadows with a similar color for each class. **b** Confusion matrix of the model for the test datasets. The values are shown as the average performance of the model over 10 independent test sets (same as **a**) with the corresponding standard deviations. The error in non-adjacent classes (e.g., normal vs. moderate) is zero for all classes (except for severe vs. mild, which has a low error of 0.01).

### 2.3 Deep inside the layers of CNN

To assess the consistency of the results, we use the Gradient-weighted Class Activation Mapping (Grad-CAM) method for saliency map visualization of the input CXRs [41]. Figures 3a-c show the original CXRs and regions of interest (ROI) labeled by our expert radiologist for three images with mild, moderate, and severe conditions; the corresponding saliency maps obtained from the AI algorithms are shown in Figs. 3d-f. All of the images have been classified correctly by our model, and the probability of the true classes are 0.96, 0.67, and 0.89, respectively. The saliency maps demonstrate consistent activation in the regions that are affected by COVID-19 pneumonia.

**Figure 3.**
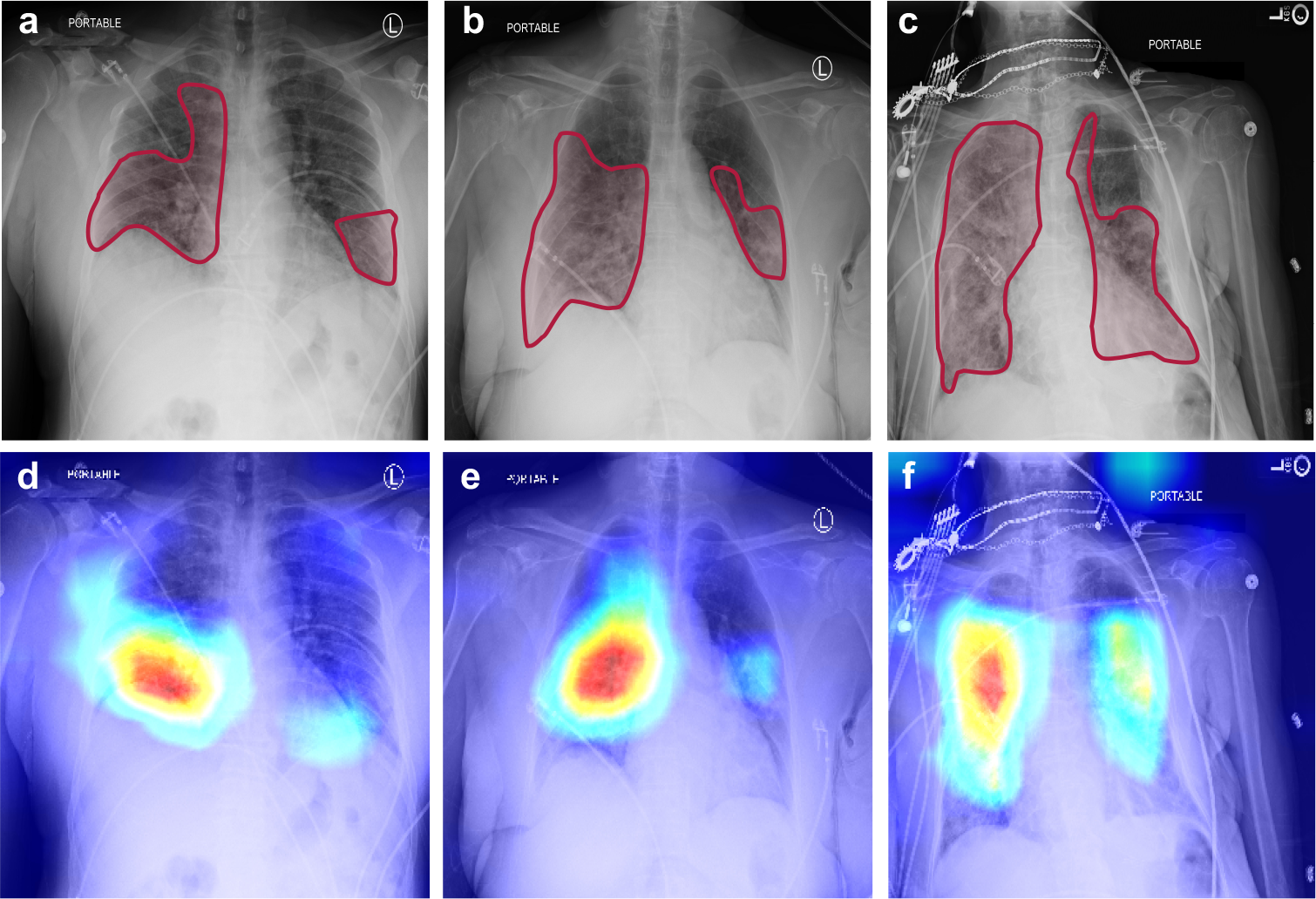
CXRs and saliency maps for three patients chosen from the unseen test dataset. The CXRs and affected areas for patients with **a** mild, **b** moderate, and **c** severe COVID-19 pneumonia (analyzed by our expert radiologist). **d-f** The corresponding saliency maps for CXRs (found by our AI approach). The true class probability for the images are 0.96, 0.67, 0.89, respectively. The saliency maps and regions of interest match well for all CXRs.

To understand the decision-making process in the CNN and visualize the extracted features, we use a pruning algorithm based on APoZ method to find the most dominant filters in each convolutional layer of the CNN. The outputs of the corresponding CNN layers under these dominant filters are shown in Figs. 4a-p for an input CXR of a patient with severe COVID-19 pneumonia. Comparing the filters to our expert radiologist’s perspective of the traditional CXRs shows that each filter highlights specific relevant parts of the CXRs similarly to the radiologist’s workflow examination. The filters from the first block of the CNN (Figs. 4a-c) focus more on highlighting different parts of the image (e.g., lung area, lower body part, etc.). In Figs. 4d-f, the filters highlight different heterogeneities of the images. Depending on the underlying anatomical structure, different parts of the CXR will have more homogenous intensities, such as the fat layers and the upper abdominal area. The aerated parts show different types of heterogeneity, caused by differences in air and fluid content. In these filters the lung fields and projected ribs are highlighted, which can help in understanding the COVID-19 disease, where we see an increased heterogeneity in the intensities of the lung fields, caused by pulmonary infiltrates. The deeper layers of the CNN extract more complex and difficult-to-interpret features (yet potentially more insightful) from the CXRs, and the final layers localize the ROIs in the image. It is clear that the large number of filters in our algorithm provides a larger range of important features and CXR areas that might offer new insights to radiologists about the disease.

**Figure 4.**
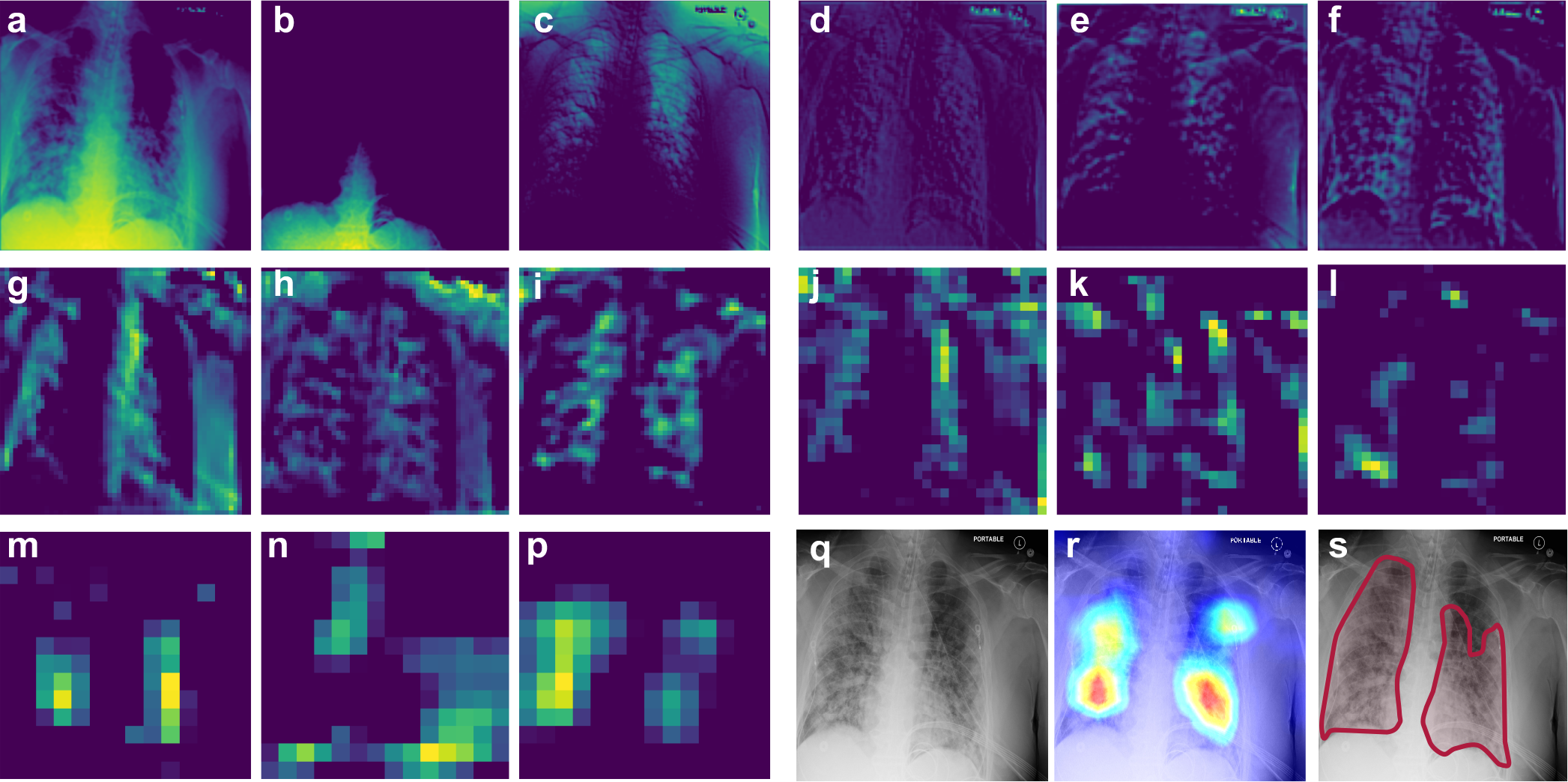
Deep inside the CNN. The representation of the dominant filters of CNN for an input CXR of a patient with severe COVID-19 pneumonia. **a-p** Outputs of the different CNN layers (from the first to the last convolutional layer) through the dominant filters. Each three filters correspond to one block of the CNN (see methods for details). **q** The original input CXR for the patient. **r** Saliency map of the input CXR. **s** Affected regions labeled by our expert radiologist.

### 2.4 Manifold learning for better CXR image visualization and analysis

We reduce the dimensionality of the output of the penultimate layer in the CNN in Fig. 1 using UMAP to visualize the distribution of the data in a two-dimensional (2D) space, called the latent space, and compare different COVID-19 severity classes (see Methods for details).

Next, we cluster the latent space using GMM into different severity regions. Fig. 5 shows the output of the model and the regions of normal, mild, moderate, and severe classes in green, blue, orange, and red colors. The results shown in Fig. 5 support the robustness of our model in classifying non-adjacent classes, as there is no overlap between their corresponding regions in the latent space. More importantly, this approach provides a visualization tool for monitoring the progression of the disease and studying the changes in the severity degree of COVID-19 for a specific patient over time. Progressions of the disease in consecutive follow-up visits for two patients are shown in Fig. 5 with their corresponding CXRs. Patient 1 is an Asian female with body mass index (BMI) of 22.5, and is in their 70’s. As it is shown in Fig. 5, the patient has normal CXR in the first visit and the severity increases in the follow-ups and unfortunately dies. The severity path and CXRs for patient 2 (African American female with BMI of 29.8, in their 60’s) are also shown in Fig. 5. This patient starts from mild stage and as the disease progresses, the CXR shows severe condition in the 5th follow-up and fortunately starts recovering in the next visits.

**Figure 5.**
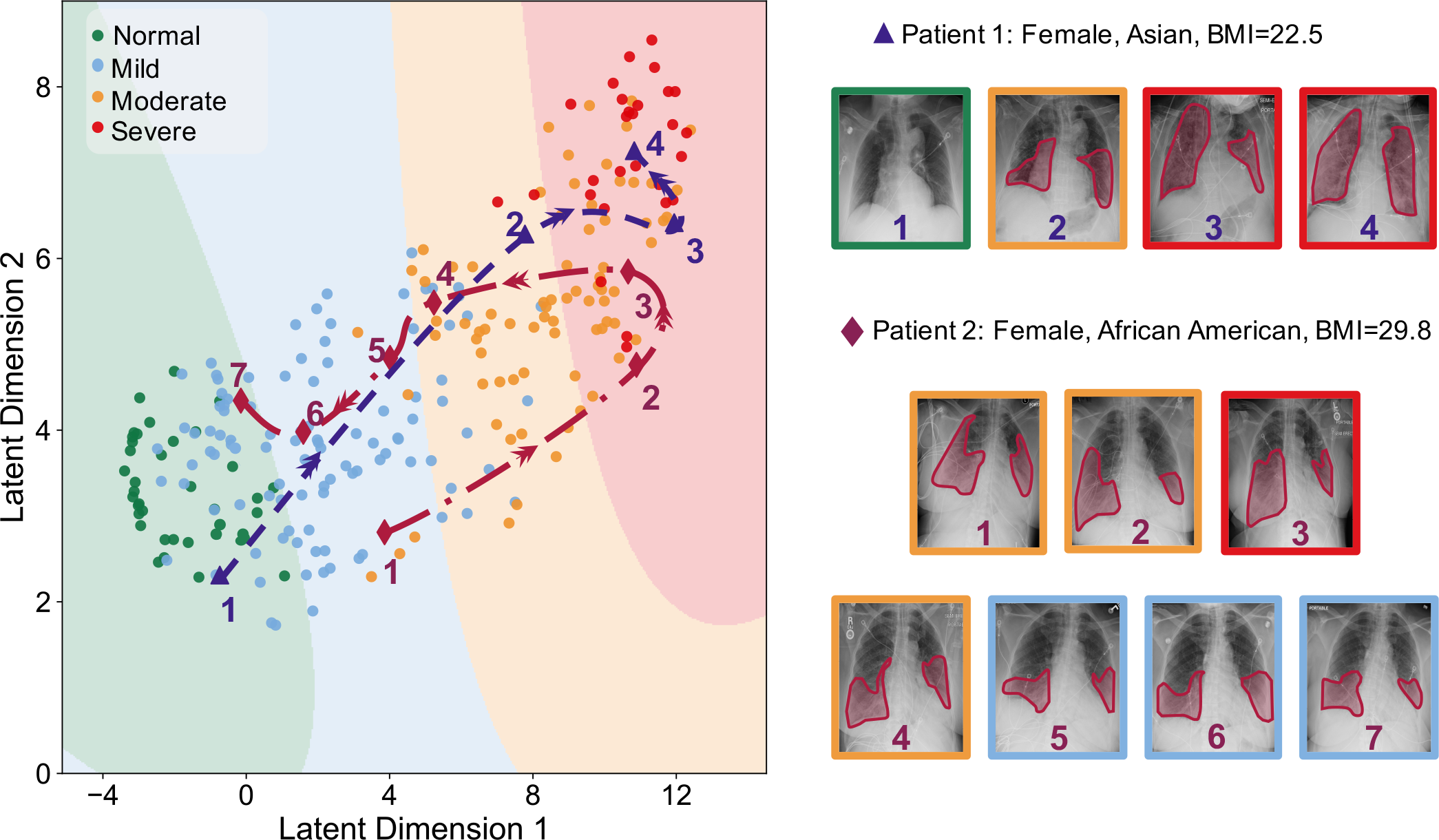
Latent space representation of the X-rays. The dimensionality of the penultimate layer of the network is reduced using UMAP, and the resulting latent space is clustered into 4 regions corresponding to different severity degrees using GMM. Green, blue, orange, and red are related to the normal, mild, moderate, and severe classes, respectively. The datapoints for each class are shown as circles with the corresponding color. The progressions of the disease over time for two patients are shown by the dashed blue and red paths in the latent space.

## 3 Discussion

With the availability of the simpler, more reliable, and faster RT-PCR test kits, the strength of AI approaches must be utilized to provide early prognostication and more subtle details about the disease than just the basic detection. With pneumonia being a catastrophic feature of COVID-19, there is an urgent need for better understanding the dynamics of the progression of the adverse effects of COVID-19 on the lung. X-ray imaging can play an important role in this endeavor as the first-line imaging tool, which is an important part of the standard protocols, requiring a low radiation dose, easy to disinfect, inexpensive, and widely available, especially in low-income/rural areas and countries.

In this study, we developed an AI system to assess the severity degree of COVID-19 pneumonia using CXRs and monitor the progression. Figures 2, 3, and 5 clearly show that our algorithm can learn to assess the severity degree of COVID-19 similar to an expert radiologist. Our algorithm can also provide more features than a typical CXR using the outputs of the CNN layers under dominant filters as shown in Fig. 4. Having these different views of a single CXR can help radiologists to better evaluate the severity of the disease. The accuracies obtained for the severity classification (Fig. 2) are good, especially knowing that the comparison is made with the labeling of only one expert radiologist. Aside from the actual classification accuracies, the separation of different severity classes in Fig. 5 clearly shows that our algorithm successfully distinguishes different classes with maximum spatial distance in the latent space between the normal and severe classes. It is also trivial why the errors in classifying mild and moderate classes are higher than those in the other two. A valuable use of the manifold-learning approach in Fig. 5 is the observation of the disease progression path for each patient. Interestingly we see that these patients take two different disease progression paths with different specifics. Patient 2, initially worsens, showing increasing signs of COVID-19-related pneumonia on sequential CXRs. However, after day 5, the patient starts recovering, showing improvement on CXRs and on day 20 is successfully discharged without re-admission. Patient 1 does not show this turning point and shows increasing disease severity on the CXRs and unfortunately dies. Understanding such turning points based on available data and relating them to the influencing factors on the disease progression are essential to create more insight into the disease trajectory to enable intelligent prediction of the disease evolution and assessment of treatment options for specific patients early on.

Having more frequent CXRs for more patients will enable the algorithm to relate the dynamics of disease progression to the clinical data and treatment details. Combined with other clinical data for a patient, CXRs can provide valuable insight into the disease progression over time and the patient’s response to treatments. This can also help us to develop a more concrete model to project at an early stage which patients might become in need for more intensive treatment and which patients are more likely to recover. AI approaches can use such data to learn these dynamics and predict the course of disease evolution for each patient. It can also be used to predict the best treatment approach at each stage of the disease. It is important to note that the applicability of our AI approach for clinical data analysis is not limited to COVID-19 and can be extended to a wide range of lung diseases. Once the algorithm is trained to learn the patterns of a disease through a series of training CXRs, the outputs of the different CNN layers under dominant filters can identify subtle patterns in the imaging data (as seen in Fig. 4 for CXRs) that can provide more detailed information than a single image. This can improve the accuracy of CXR reading by a radiologist. In a bigger picture, our algorithm can be adopted and extended to include other forms of imaging data for a wider range of medical diagnosis.

## 4 Methods

### 4.1 COVID-19 dataset population and labeling procedure

Patients included retrospectively for this study are selected as a consecutive sample who had a clinically performed positive COVID-19 RT-PCR test related to their CXR imaging date at Emory Healthcare affiliated hospitals (Atlanta, GA, USA). The CXRs are collected from January 1st, 2020 to May 1st, 2020, and the need for informed consent is waived by the institutional review board (IRB). Both in-patients and out-patients are included. All CXRs during the entire hospital admission of the patient are collected, ranging from 1–30 images. CXRs are acquired in the posteroanterior (PA) or anteroposterior (AP) projection according to standard clinical protocols. Lateral X-rays were present for some patients, however not used for the current study. Depending on patient status, CXRs are taken with a portable X-ray unit and in a supine, erect, or semi-erect position. All CXRs have been labeled by an expert cardiothoracic radiologist with 15+ years of experience reading CXRs. All the images are blinded and randomized for the qualification of disease severity, and the reader had no insight into clinical data and/or outcomes. The CXRs are classified into normal, mild, moderate, and severe categories. This is done based on clinical experience of the reader in addition to the following guidelines:

#### Normal

no opacities and/or abnormalities, indicative of pneumonia, are present.

#### Mild

less than 50% of the lung area is affected by pneumonia-related abnormalities. Patchy (partly peripheral) opacities are present in the lower and mid lung zones

#### Moderate

approximately 50% of the lung area is affected by pneumonia-related abnormalities. Opacities are present, often bilateral, in the mid and lower zones.

#### Severe

more than 50% of the lung area is affected by pneumonia-related abnormalities. Opacities are dense, often bilateral, and can affect all lung zones (lower, mid, and upper).

In addition to the images, basic demographics are collected such as age (at the scan time), body-mass index (BMI), and gender. All images are de-identified according to Health Insurance Portability and Accountability Act (HIPAA) and hospital-specific guidelines. Patients with an age above 90 were noted as > 90. DICOM headers are anonymized and only contain technique-related information, such as information about position of the X-ray (supine, semi-erect etc.) and whether it is a portable X-ray machine or not; all patient-related data are removed. Patients and imaging dates are coded to keep the longitudinal information between CXRs from the same patient. In addition, the date and time stamp, burned into the CXRs, are removed.

### 4.2 DL-based severity classification model

We divide the dataset into 80% training and 20% test sets per class and use the bootstrap sampling method to create 10 different sets of training and test data to independently train and evaluate the performance of the model. Due to the limited number of CXRs, we use a two-stage transfer-learning approach for training the DL-based method for predicting the severity of COVID-19. We first use the pre-trained convolutional layers of the VGG-16 network for the classification task over the Radiological Society of North America (RSNA) pneumonia dataset that includes 25684 CXRs corresponding to normal, lung opacity, and other classes of lung pathologies (e.g., pulmonary edema, atelectasis, lung cancer, etc.). Then, we transfer the fine-tuned convolutional layers of the network and add fully connected layers to the transferred network, trained by our CXRs, for classifying the severity of COVID-19.

All the CXR images are resized to 224 × 224 and preprocessed to scale the pixels between -1 and 1. We use random rotations, horizontal and vertical shifts, random shears, and random zooms to augment the training set for better generalization. The network includes 13 convolutional layers (5 blocks of convolutional layers, each one having 2, 2, 3, 3, and 3 convolutional layers, respectively) transferred from VGG-16 [50] followed by an average pooling layer, a flatten layer, a dense layer of 256 neurons with the rectified linear unit (ReLU) activation function and the output layer with 4 neurons and softmax activation function. We use a categorical cross-entropy loss function and 1.5, 2, 2, 3 as the cost of error on each class (normal, mild, moderate, and severe, respectively) to handle the unbalanced dataset.

To find the regions of interest for the CNN, we use the Grad-CAM method. The output of our severity classification model will be the saliency map of the input CXR and the class probabilities. Algorithms are implemented in Python using the Keras package and trained on a workstation with an NVIDIA RTX2080 GPU, a Core i7 CPU, and 32 GB of RAM.

### 4.3 Filter representation model

To understand the role of convolutional layers in the CNN and the details of the decision-making process, we visualize the outputs of each layer of the CNN under different applied filters. We first find the most dominant filters by applying the APoZ pruning method over the training dataset. We keep 1% of the filters at each CNN layer with highest average activity (i.e., *l*_0_-norm) over the training set and visualize the corresponding outputs of those CNN layers for any given input CXR.

### 4.4 Dimensionality reduction and latent space representation model

The output of the penultimate layer (i.e., the last fully connected layer before the output layer of the CNN) is used for the latent space representation. We reduce the dimensionality of the data from 256 to 2. The number of neighbors in the UMAP algorithm are 5; the distance measure is correlation; and the minimum distance is 0.3. The UMAP is trained using the same training dataset on which the CNN is trained; none of the test samples are used during the training stage of the latent space representation model. After reducing the dimensionality to 2, we train a GMM to cluster the latent space into the regions for normal, mild, moderate, and severe COVID-19 pneumonia. The number of Gaussians (i.e. the constituents of the mixture in the GMM that model the distribution of the data) is selected as 4, and the labels are imposed as colors to specify the regions after training.

### Compliance

All the approaches and methods used the anonymized and de-identified CXR data according to the guidelines of Emory University and Georgia Tech and approved by the IRB. All procedures followed HIPAA guidelines. The Emory IRB approved the collection of this data and waived the need of informed consent. The part of the research that is performed at Georgia Tech was based on totally de-identified data and the need for IRB is waived by Georgia Tech IRB.

## Supporting information

Monitoring severity of the disease for a patient.

## Data Availability

The dataset that is used for the first stage of the transfer learning during the training is publicly available on https://www.kaggle.com/c/rsna-pneumonia-detection-challenge/data. The datasets generated during and/or analyzed (i.e., Emory COVID-19 CXR dataset) during the current study are available upon reasonable request. For more information, please contact Dr. Carlo De Cecco and Dr. Ali Adibi.

